# Longitudinal monitoring of DNA viral loads in transplant patients using quantitative metagenomic next-generation sequencing

**DOI:** 10.1101/2021.07.14.21260507

**Authors:** Ellen C. Carbo, Anne Russcher, Margriet E.M. Kraakman, Caroline S. de Brouwer, Igor A. Sidorov, Mariet C.W. Feltkamp, Aloys C.M. Kroes, Eric C.J. Claas, Jutte J.C. de Vries

## Abstract

**Introduction:** Immunocompromised patients are prone to reactivations of multiple latent DNA viruses. Viral load monitoring by single-target quantitative PCRs (qPCR) is the current cornerstone for virus quantification. In this study, a metagenomic next-generation sequencing (mNGS) approach was used for identification and load monitoring of transplantation-related DNA viruses.

**Methods:** Longitudinal plasma samples from six patients that were qPCR-positive for cytomegalovirus (CMV), Epstein-Barr virus (EBV), BK polyoma virus (BKV, adenovirus (ADV), parvovirus B19 (B19V), and torque teno-virus (TTV) were sequenced using the quantitative metagenomic Galileo Viral Panel Solution (Arc Bio, LLC) reagents and bioinformatics pipeline combination. Qualitative and quantitative performance was analysed with focus on viral load ranges relevant for clinical decision-making.

**Results:** All pathogens identified by qPCR were also identified by mNGS. In addition, BKV, CMV, and HHV6B were detected by mNGS which were not ordered initially but could be confirmed by qPCR. Viral loads determined by mNGS correlated with the qPCR results, with inter-method differences in viral load per virus ranging from 0.19 log_10_ IU/ml for EBV to 0.90 log_10_ copies/ml for ADV. TTV, analysed by mNGS in a semi-quantitative way, showed a mean difference of 3.0 log_10_ copies/ml. Trends over time in viral load determined by mNGS and qPCR were comparable, and clinical thresholds for initiation of treatment were equally indicated by mNGS.

**Conclusion:** The Galileo Viral Panel for quantitative mNGS performed comparable to qPCR with regard to detection and viral load determination, within clinically relevant ranges of patient management algorithms.

## Introduction

Opportunistic viral infections frequently occur after solid organ or hematopoietic cell transplantation, with associated morbidity and mortality of up to 40% [1]. Successful prevention and early detection of viral infections including reactivations are the cornerstone of transplant patient management. For effective pre-emptive and therapeutic treatment strategies, accurate viral load quantification is essential. Typically, in immunocompromised hosts, multiple viruses can reactivate simultaneously, which makes comprehensive identification of replicating pathogenic viruses essential. Monitoring of opportunistic viral infections in transplant patients currently most frequently is performed by multiple single-plex quantitative PCRs.

Metagenomic next-generation sequencing (mNGS) is increasingly being applied for the identification of pathogens in undiagnosed cases suspected of an infectious disease [2][3][4]. Quantification of viral loads by means of mNGS remains a challenge [5][6][7][8]. Complicating factors are the varying amount of background sequences from host and bacterial origin, technical bias affecting target sequence depth, unselective attribution of reads, and the amount of calibration curves that are needed simultaneously when using untargeted sequencing for viral load calculations. Reports comparing mNGS with qPCR showed correlation with normalized sequence read counts but never as accurate as qPCR for viral load prediction[5]. Other previous research with regard to quantification of shotgun sequence read counts focussed mainly on differential expression of RNA [9][10][11][12]. Recently, the Galileo Viral Panel (Arc Bio, LLC) has been designed as a quantitative mNGS approach for ten transplant-related DNA viruses [13][14]. This all-inclusive approach encompasses the library preparation kit, controls, calibration reagents, and cloud-based user-friendly software for bioinformatic analysis. Previous data on the performance of this mNGS approach showed that the analytical performance was comparable to qPCR results with regard to the limits of detection, limits of quantification, and inter-assay variation [13][14].

In this study, we analysed the performance of the Galileo Viral Panel for longitudinal viral load quantification in transplant patients over time. Subsequent samples from six transplant patients with proven infections or reactivations with transplantation-related DNA viruses (adenovirus, ADV, BK polyomavirus, BKV, cytomegalovirus, CMV, Epstein-Barr virus, EBV, human herpes virus type 6A, HHV-6A, human herpes virus type 6B, herpes simplex type 1, HSV-1, herpes simplex type 2, JC polyomavirus, JC polyomavirus, JCV, varicella zoster virus, VZV, parvovirus B19, B19V, and torque teno virus, TTV) were analysed in comparison with qPCR. Accuracy of viral load quantification by mNGS was studied in relation to thresholds that had been used for the initiation of treatment.

Furthermore, we investigated the additional detection of DNA viruses identified by the broad mNGS approach, for which initially no targeted qPCR had been ordered.

## Methods

### Patients and sample selection

Six immunocompromised patients (one allogeneic stem cell transplant patient, four kidney transplant patients, one haematological patient) were selected based on available follow-up EDTA plasma samples that previously tested positive for one or more transplantation-related DNA viruses. Samples had been previously (July 2008 – December 2019) sent to the Clinical Microbiological Laboratory (CML) of the Leiden University Medical Center (LUMC, The Netherlands) for viral load monitoring. The samples were respectively qPCR positive for ADV, BKV, CMV, EBV, B19V, and TTV, with a wide range of viral loads. Patient plasma samples were stored at -80°C until mNGS analysis.

### Ethical approval

Approval was obtained from the ethical committee from the LUMC (P11.165 NL 37682.058.11, and Biobank Infectious Diseases protocol 2020-03 & 2020-04 B20.002).

### Extraction of nucleic acids, internal controls

Patient plasma samples were spiked with an internal control (baculovirus, Arc Bio, LLC) prior to extraction. Nucleic acids were extracted from 200 μl plasma using the MagNApure 96 DNA and Viral NA Small volume extraction kit on the MagNAPure 96 system (Roche Diagnostics, Almere, The Netherlands) with 100 μL output eluate. The eluate was concentrated using vacuum centrifugation by a SpeedVac vacuum concentrator (Thermo Scientific) to a volume of 26 µl.

### Library preparation and sequencing

Sequence libraries were prepared using the Galileo Viral Panel sequencing kit (Arc Bio, LLC., Cambridge, MA, USA) according to the manufacturer’s instructions. The protocol is based on enzymatic fragmentation at 37°C for 5 minutes, followed by end repair and A-tailing at 65°C for 30 minutes. Subsequently, fragments were ligated using unique dual-index adapters at 20°C for 15 min and purified using magnetic Kapa Pure Beads (Roche). Human DNA was depleted using human depletion reagents at 45°C for 2 hours followed by 45 °C for 15 minutes, after which libraries were amplified using library amplification primers for 45 °C for 30 seconds, by 14 cycles of 98°C for 10 seconds and 65°C for 75 seconds and 65°C for 5 minutes. The final library preparation products were purified using magnetic Kapa Pure Beads (Roche) and quantified using a Qubit fluorometer (Thermo Fisher) followed by equally pooling using the Arc Bio calculation pooling tool. After a final quantity and quality check using a Bioanalyzer (Agilent), samples were sequenced using the NovaSeq6000 sequencing system (Illumina, San Diego, CA, USA) at GenomeScan B.V. (Leiden, the Netherlands) aiming at 10 million reads per library.

### Calibration samples

Initial calibration runs were performed testing the multi-analyte mixture (MAM) of whole-virus particles at viral loads of 0, 1,000, 5,000, 10,000, and 100,000 copies/ml or IU/ml plasma, in quintuple (Arc Bio, LLC) for the following 10 viruses: ADV, BKV, CMV, EBV, HHV-6A, HHV6B, HSV-1, HSV-2, JCV, and VZV. For TTV and B19V, no Arc Bio calibrator panels were available, and therefore the Galileo Signal values were plotted against the calibrator plot of other viruses that showed optimal agreement with the viral load (respectively JCV and VZV), representing a semi-quantitative result.

### Bioinformatic analysis

After demultiplexing of the sequence reads using bcl2fastq (version), FASTQ files were uploaded to the Galileo Analytics web application [13][15] which automatically processes data for quality assessment and pathogen detection using a custom database of DNA viruses involved in transplant-associated infections: ADV, CMV, EBV, HHV-6A, HHV-6B, HSV-1, HSV-2, JCV, VZV, B19V and TTV.

Galileo Analytics web application aligns sequence reads to the genomes of the DNA viruses in their calibration kit and scores these read alignments based on complexity, uniqueness and alignment scores and reports this in a signal value. The signal value is normalized for read counts across libraries, correcting for differences in genome lengths and technical bias, based on the spiked-in normalization controls. The Signals reported are related to the genomic depth and the observed amount of viral DNA being present in a sample, belonging to non-confounding genomic regions [13]. The sample Signals were visualized in linear calibration curves (**Supplementary Figure 1**).

### Analysis of performance, and additional findings

Performance of the metagenomic Galileo Viral Panel assay was assessed in comparison with routine qPCR, analyzing both qualitative and quantitative detection. Additional findings by mNGS were confirmed by additional qPCR analysis. In case no remaining sample was available, the Galileo Analytics software results were compared with results from analysis using alternative bioinformatic tools: metagenomic taxonomic classifier Centrifuge (1.0.4-beta) [16] and *de novo* assembly-based viral metagenomic analysis software Genome Detective [17].

## Results

### Calibration curves

After metagenomic sequencing, the viral loads were calculated for each virus by the Galileo Analytics web application. Signals of both the calibrators and patient plasma samples were plotted in load graphs (**Supplementary Figure 1**) and the corresponding viral load of the patient samples was extrapolated. Since no calibrator panels for B19V and TTV virus were available, these signals were plotted against other calibration curves of viruses that showed the optimal agreement with the known viral load for semi-quantitative detection. All calibration sample signals correlated well with the titer (R^2^ range 0.84-0.92).

### Viral load by mNGS versus qPCR

In total six patients were tested by qPCR and mNGS for quantification of different viruses at subsequent time points. Agreement between the methods for qualitative detection was 100% for the viruses targeted by PCR. Quantitative results per patient are shown in **Table 1**, and **Figure 1** depicts viral loads by mNGS versus qPCR per target virus. CMV and EBV viral loads showed highest agreement, with a maximum difference in viral load of 0.70 log_10_ IU/ml. Mean differences in viral loads were 0.43 for CMV and 0.19 log_10_ IU/ml for EBV. For ADV, viral loads were higher when quantified with mNGS with a mean difference of 0.90 log_10_ c/ml. For BKV, viral loads by mNGS were lower in comparison with qPCR, with a mean difference of 1.32 log_10_ c/ml. When taking into account viral loads measured above the limit of quantification of 2.5 log_10_ c/ml, as applied in our diagnostic qPCR for BKV, the mean difference is 0.62 log_10_ c/ml and a trend towards better agreement with higher viral loads could be observed. Semi-quantitative detection of B19V and TTV viruses by mNGS resulted in mean differences of, respectively, 0.39 log_10_ IU/ml and 3.0 log_10_ c/ml in comparison with qPCR.

**Table 1.** Viral load quantification by qPCR and mGNS per patient sample.

| Patient-sample | Virus | Viral load qPCR | Viral load qPCR (log<sub>10</sub>) | Viral load mNGS | Viral load mNGS (log<sub>10</sub>) | ΔqPCR-mNGS (log<sub>10</sub>) |
| --- | --- | --- | --- | --- | --- | --- |
| P1-S1 | ADV | 675 c/mL | 2,83 c/mL | 1277 c/mL | 3,11 c/mL | 0,28 c/mL |
| P1-S2 |  | 4517 | 3,65 | 66273 | 4,82 | 1,17 |
| P1-S3 |  | 34740 | 4,54 | 287844 | 5,46 | 0,92 |
| P1-S4 |  | 136900 | 5,14 | 1435130 | 6,16 | 1,02 |
| P1-S5 |  | 60540 | 4,78 | 777172 | 5,89 | 1,11 |
| P2-S1 | BKV | 796 c/mL | 2,90 c/mL | 3 c/mL | 0,48 c/mL | -2,42 c/mL |
| P2-S2 |  | 614 | 2,79 | 3 | 0,48 | -2,31 |
| P3-S3 |  | 233700 | 5,37 | 9011 | 3,95 | -1,41 |
| P4-S4 |  | 2401000 | 6,38 | 1857785 | 6,27 | -0,11 |
| P5-S5 |  | 71480 | 4,85 | 32321 | 4,51 | -0,34 |
| P3-S1 | CMV | 2370 IU/mL | 3,37 IU/mL | 6246 IU/mL | 3,80 IU/mL | 0,42 IU/mL |
| P3-S2 |  | 122800 | 5,09 | 275657 | 5,44 | 0,35 |
| P3-S3 |  | 10680 | 4,03 | 22242 | 4,35 | 0,32 |
| P3-S4 |  | 4915 | 3,69 | 11366 | 4,06 | 0,36 |
| P3-S5 |  | 9156 | 3,96 | 46231 | 4,66 | 0,70 |
| P3-S1 | EBV | 2083 IU/mL | 3,32 IU/mL | 4581 IU/mL | 3,66 IU/mL | 0,34 IU/mL |
| P3-S2 |  | 12970 | 4,11 | 1573 | 4,20 | 0,09 |
| P3-S3 |  | 17710 | 4,25 | 14549 | 4,16 | -0,09 |
| P3-S4 |  | 10500 | 4,02 | 15077 | 4,18 | 0,16 |
| P3-S5 |  | 7723 | 3,89 | 14844 | 4,17 | 0,28 |
| P4-S1 | TTV* | 140 c/mL | 2,15 c/mL | 4 c/mL | 0,60 c/mL | -1,54 c/mL |
| P4-S2 |  | 2400000 | 6,38 | 5142 | 3,71 | -2,67 |
| P4-S3 |  | 5,7E+09 | 9,76 | 319074 | 5,50 | -4,25 |
| P4-S4 |  | 2,4E+08 | 8,38 | 46261 | 4,67 | -3,71 |
| P5-S1 | B19V* | 1,34E+11 IU/mL | 11,13 IU/mL | 2,07E+11 IU/mL | 11,32 IU/mL | 0,19 IU/mL |
| P5-S2 |  | 1407365 | 6,15 | 1235416 | 6,09 | -0,06 |
| P5-S3 |  | 45846 | 4,66 | 41787 | 4,62 | -0,04 |
| P6-S1 | B19V* | 4,07E+10 IU/mL | 10,61 IU/mL | 4,37E+11 IU/mL | 11,64 IU/mL | 1,03 IU/mL |
| P6-S2 |  | 5309308 | 6,73 | 9376953 | 6,97 | 0,25 |
| P6-S3 |  | 8569 | 3,93 | 49601 | 4,70 | 0,76 |

\*B19V and TTV results were considered semi-quantitative since no Arc Bio calibration samples were available for these targets.

**Figure 1.**
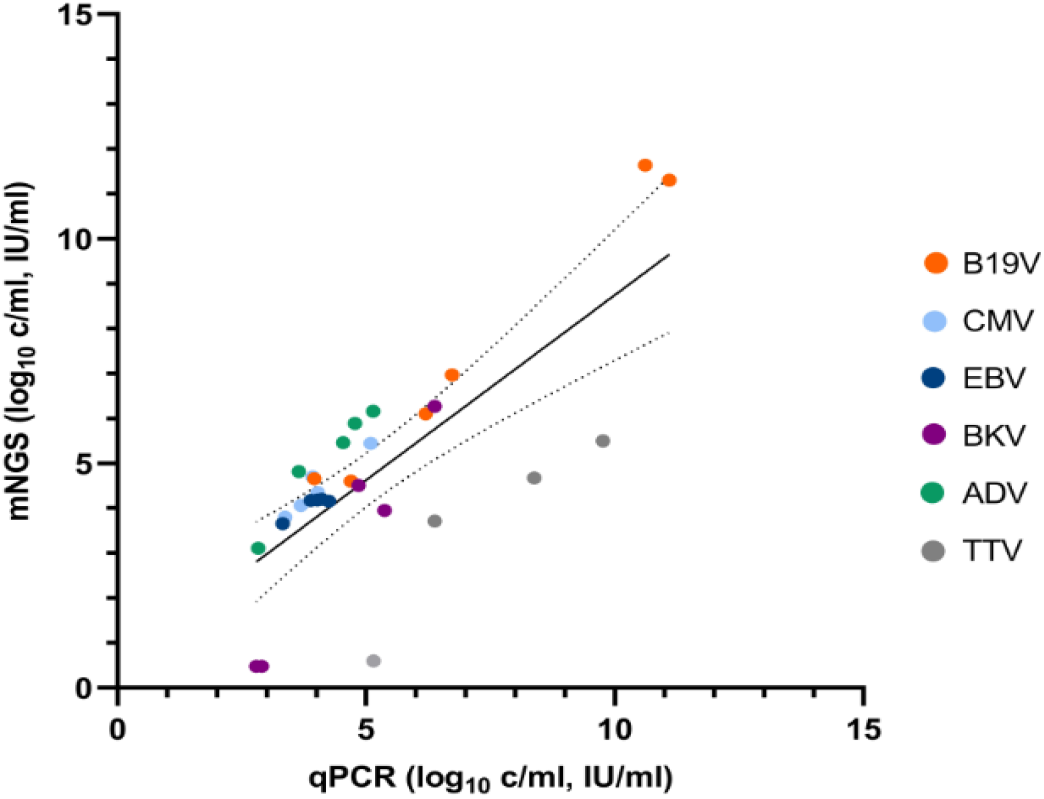
Viral loads as predicted by Galileo Viral Panel mNGS versus qPCR (copies/ml for ADV, BK, and TTV, and IU/ml for CMV, EBV and B19V). B19V and TTV results were considered semi-quantitative since no Galileo calibration panels were available for these targets.

### Longitudinal patient follow-up and clinical decision-making

Furthermore, for each patient the viral loads over time were plotted in graphs with clinical information about symptomatology and treatment (**Figure 2**). For CMV, EBV and BKV, in our clinical practice, specific viral load thresholds are used to decide whether immunosuppression should be tapered and/or antiviral therapy should be administered. Viral load quantification around these thresholds showed good agreement in identifying these clinical decision-making breakpoints. In Patient 3, antiviral treatment with foscarnet was started for CMV-reactivation when viral load measured by qPCR exceeded 4.0 log_10_ IU/ml. By mNGS, this critical threshold for treatment initiation was correctly identified with a viral load by mNGS of 5.44 log_10_ IU/ml. In the same patient, rituximab was administered when the EBV load by qPCR was repeatedly above the threshold of 4.0 log_10_ IU/ml, consistently quantified thrice above 4.0 log_10_ IU/ml before administration of rituximab both by qPCR and mNGS.

**Figure 2.**
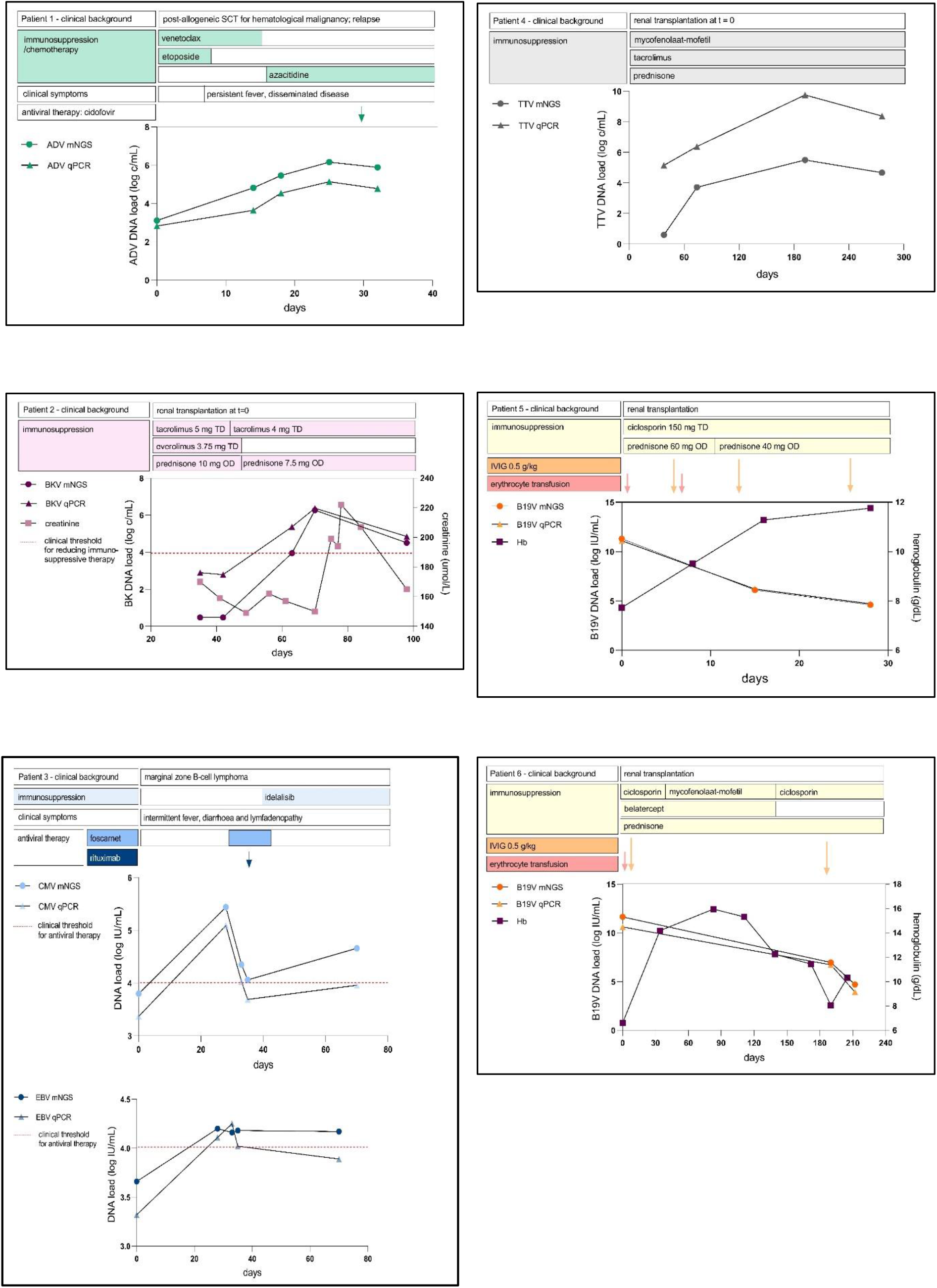
Longitudinal follow-up of DNA viral loads in immunosuppressed patients over time, as predicted by mNGS (Galileo Viral Panel, Arc Bio) versus qPCR. Clinical information and therapeutic agents are included.

For B19V, ADV and TTV, no predefined thresholds were used for changing the treatment regimen. For all viruses, the observed trends in load over time in each patient were comparable for qPCR and mNGS, despite the semi-quantitative nature of the B19V mNGS assay. Effect of treatment (anti-viral drugs, immunoglobulins and/or tapering of immunosuppressive drugs) in patients was estimated by follow-up of viral loads by qPCR. For B19V in Patient 5 and 6, the effect of intravenous immunoglobulins (IVIG) could be assessed by the decreasing viral load in the weeks after administration, as observed by mNGS. For ADV, in patient 1, antiviral therapy with cidofovir was started when increasing viral loads were consistently detected, both by qPCR and mNGS.

### Additional findings

For some samples additional viral reads were detected in the pathogenic mNGS reports, of viruses that were not initially tested for by qPCR (**Supplementary Table 1**). Most additional findings were supported by a secondary bioinformatic analysis using Centrifuge and Genome Detective: BK (1 patient), CMV (1 patient), HHV-6B (1 patient), and TTV (4 patients). In a few cases, additional findings were not confirmed by a second analysis leaving some low mNGS signals for CMV, EBV and HSV. JCV was detected by mNGS in a sample with a high concentration of BKV, possibly indicating forced alignment contamination due to high sequence homology between JCV and BKV [13][14].

## Discussion

In this study, the performance of a quantitative mNGS assay for longitudinal follow-up of DNA viral loads was analysed in six immunocompromised patients. Viral loads determined by mNGS were comparable with loads determined by qPCR, and differed less than 1 log_10_ for DNA viruses with calibration panels available, in line with previous studies [13][14]. In the current study, the performance of viral loads assessed by mNGS was also evaluated with regard to clinical decision-making. In the management of reactivating viruses in immunocompromised patients, local and international guidelines use viral load breakpoints to decide whether antiviral therapy should be administered or whether immunosuppression should be tapered [18][19][20][21][22]. When local clinical breakpoints were considered for each virus, mNGS performed comparable to qPCR to identify the clinically relevant breakpoints. B19V is not considered to be a reactivating virus, but quantification may be helpful to distinguish clinically relevant replicative infection from merely DNA remnants [23]. In the range of these breakpoints, viral loads were adequately determined by mNGS to guide clinical decision making. Additionally, the longitudinal trend was similar in comparison with qPCR, indicating precision of mNGS for clinical quantification and reliable indication of the trend in viral load. Clinical decision-making is often guided by follow-up of viral load trends, in addition to the cross-sectional viral load measurements for viral infections without available thresholds.

The principle of a quantitative catch-all approach to detect all transplantation-related viruses in a single run is an attractive feature in the clinical follow-up of the immunocompromised host.

Simultaneous reactivation of latent viruses during immunocompromised episodes is common. Co-infection rates of up to 32% have been described using PCR and, importantly, were associated with higher rates of acute rejection or graft dysfunction[24]. Co-infections may be missed when ordering targeted PCRs, while the catch-all approach of mNGS could guarantee that active infections are not overlooked. Indeed, our approach showed a complementary yield of seven reactivating viruses in five patients, which had not been identified earlier by qPCR. Some of these unnoted viruses are not considered pathogenic, like TTV, which merely reflects the level of functional immunity and could serve as a marker for balancing immunosuppressive treatment [25][26][27]. A significant complementary virus identification yield by mNGS in transplant patients of 31/49 plasma samples was also reported by Sam et al [14], with the majority being viruses considered pathogenic. These findings show that mNGS could improve pathogen detection in clinical practice.

Another advantage of mNGS would be its capacity to genotype viruses and detect mutations associated with antiviral resistance, without the need for additional, time consuming, target-specific ‘wet’ lab procedures that could delay diagnosis and treatment. As an example, Patient 3 in our study was treated with foscarnet for persistent CMV reactivation pending the results of mutational analysis after clinical failure of valganciclovir treatment. If the results of mutational analysis had been immediately available, resorting to second-line treatment may have been avoided.

Widespread implementation of mNGS approaches in clinical diagnostic settings has been limited by several factors. The ‘wet’ lab protocols can be time-consuming, costly, and have a relatively long turnaround time mainly due to the time required for sequencing. With various sequencing techniques still rapidly evolving, the costs and sequencing turnaround time of such protocols is expected to improve considerable in the future [28]. Furthermore, bioinformatic skills are generally needed for validation and implementation as a diagnostic assay. User-friendly, all-in one mNGS data-analysis software packages for cloud-based, automated analysis, enables use in laboratories with minimal bioinformatic knowledge and high-performance computing capacity.

Limitations in this current study are the relatively low number of samples and viruses when considering a metagenomic approach, including two viruses without calibration panels available. This small-scale study provides a proof-of-principle demonstration in a retrospective design that the current version of the Research Use Only Galileo Viral Panel enables longitudinal viral load monitoring by mNGS. It is expected that after these initial studies indicating high performance in terms of limit of detection and quantification, inter-run precision and prospective viral load monitoring, the kit and software will be expanded to include more viruses, calibration samples and potentially fit for different sample types. Furthermore, technical and bioinformatic features might be evolved in future versions of the assay.

Overall, viral metagenomic sequencing is a promising approach not only DNA virus detection and identification, but also for reliable estimation of the viral load in a clinical setting, and potentially mutational typing for drug sensitivity analysis. Several milestones essential for implementation in diagnostics settings have been met by the specific assay used in this study: the limits of detection, the limits of quantification, precision and overall technical performance, which were comparable with qPCR assays. Precise quantification was accomplished by read normalization based on a designed control. These accomplishments pave the way for further developments and optimization of quantitative metagenomic sequencing for longitudinal viral load monitoring and beyond.

## Data Availability

N/A

## Acknowledgements

We thank Gavin Wall, Naissan Hussainzada, and Meredith Carpenter (Arc Bio) for their technical and logistic support.

---

**Supplementary Figure 1.**
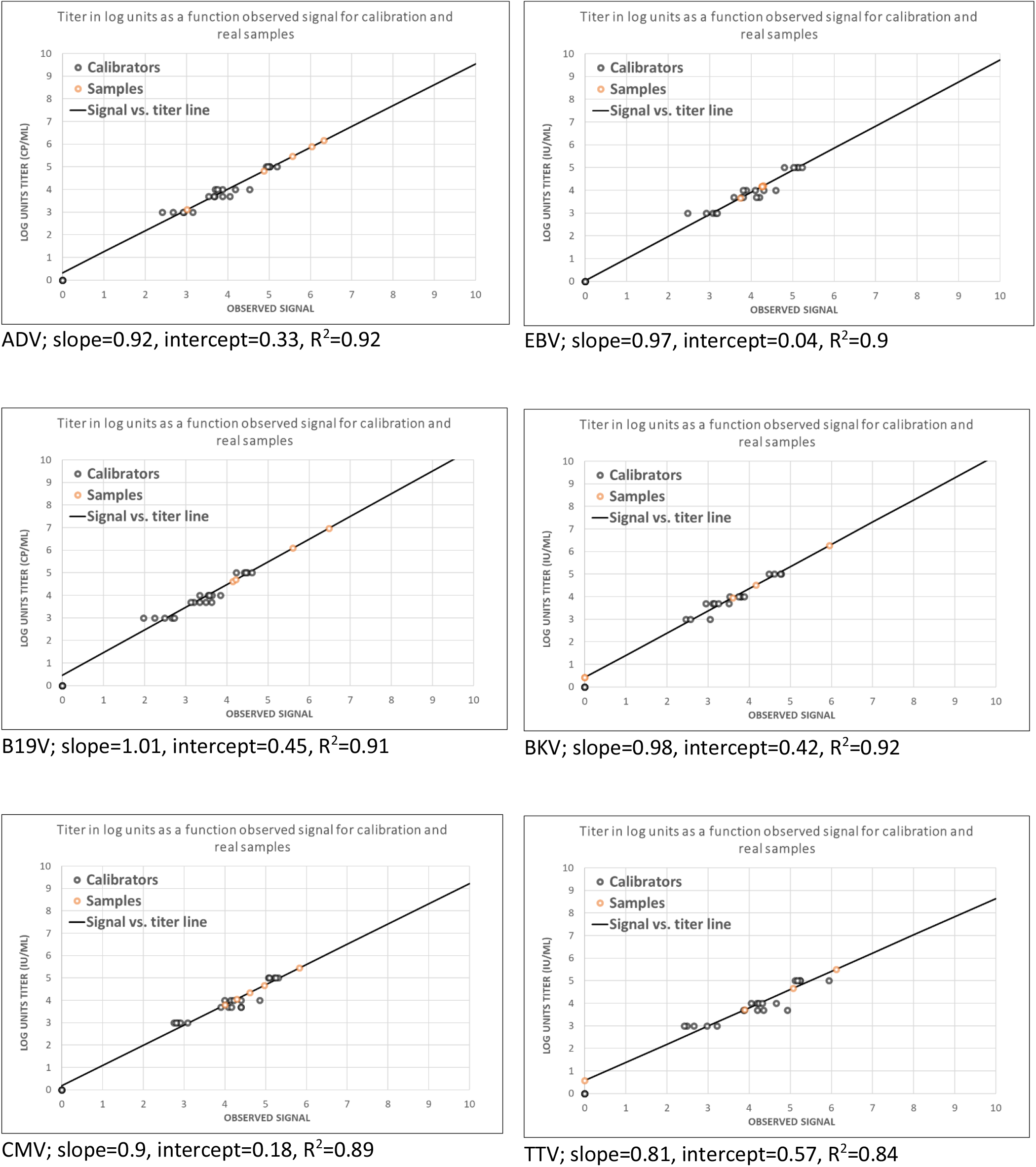
Calibration graphs of the six viruses in six patients in this study with associated slope, intercepts and R^2^ values. Concentrations are expressed in log_10_ copies or IU/ml. Calibrator samples are shown in black dots, clinical samples in orange.

**Supplementary Table 1.** Additional findings of the metagenomic Galileo Viral Panel compared to Centrifuge and Genome Detective software. For Galileo Analytics, results are presented as viral load in log10 c/mL or IU/ml. For Centrifuge and Genome Detective, results are presented as absolute amount of reads classified per species or genus. For TTV reads during centrifuge analysis, Anelloviridae reads are shown.

| P1<br>ADV |  | S1 | S2 | S3 | S4 | S5 |
| --- | --- | --- | --- | --- | --- | --- |
| BKV | Galileo Analytics |  | 3.67 | 4.03 | 3.78 |  |
|  | Centrifuge |  | 47 | 74 | 67 |  |
|  | Genome Detective |  | 84 | 113 | 35 |  |
| CMV | Galileo Analytics | 2.45 | 3.69 | 3.84 | 3.84 | 3.16 |
|  | Centrifuge | 18 | 123 | 83 | 53 | 1 |
|  | Genome Detective | 0 | 12 | 156 | 2 | 0 |
| EBV | Galileo Analytics |  | 1.22 |  |  |  |
|  | Centrifuge |  | 0 |  |  |  |
|  | Genome Detective |  | 0 |  |  |  |
| B19V | Galileo Analytics | 3.85 | 3.65 |  | 3.55 |  |
|  | Centrifuge | 0 (1 AADPA<sup>†</sup>) | 2 (147 AADPA) |  | 0 (1 AADPA) |  |
|  | Genome Detective | 0 | (140 AAV2<sup>‡</sup>) |  | (124 AAV2) |  |
| TTV | Galileo Analytics | 5.54 | 6.18 | 4.98 | 4.61 | 4.39 |
|  | Centrifuge | 2382 | 3525 | 489 | 137 | 2 |
|  | Genome Detective | 11426 | 22595 | 2719 | 823 | 2 |

<sup>†</sup> AADPA = Adeno-associated dependoparvovirus A; <sup>‡</sup>AAV2 = adeno-associated virus 2

| P2 BKV |  | S1 | S2 | S3 | S4 | S5 |
| --- | --- | --- | --- | --- | --- | --- |
| ADV | Galileo Analytics | 2.94 |  | 2.94 |  | 2.44 |
|  | Centrifuge | 3 |  | 0 |  | 1 |
|  | Genome Detective | 0 |  | 0 |  | 0 |
| CMV | Galileo Analytics |  | 2.6 | 4.46 | 5.07 |  |
|  | Centrifuge |  | 13 | 279 | 1056 |  |
|  | Genome Detective |  |  | 110 | 975 |  |
| TTV | Galileo Analytics | 3.87 | 4.84 | 5.54 | 4.71 | 4.99 |
|  | Centrifuge | 96 | 1196 | 953 | 56 | 1511 |
|  | Genome Detective | 406 | 7037 | 5900 | 241 | 11141 |
| VZV | Galileo Analytics |  |  | 2.3 |  |  |
|  | Centrifuge |  |  | 1 |  |  |
|  | Genome Detective |  |  | 0 |  |  |
| JCV | Galileo Analytics |  |  |  | 4.06 |  |
|  | Centrifuge |  |  |  | 0 |  |
|  | Genome Detective |  |  |  | 0 |  |
| HSV | Galileo Analytics |  | 0.87 |  |  |  |
|  | Centrifuge |  | 1 |  |  |  |
|  | Genome Detective |  |  |  |  |  |

| P3<br>CMV/EBV |  | S1 | S2 | S3 | S4 | S5 |
| --- | --- | --- | --- | --- | --- | --- |
| BKV | Galileo Analytics |  | 4.99 | 4.98 | 4.89 | 5.12 |
|  | Centrifuge |  | 22 | 60 | 33 | 21 |
|  | Genome Detective |  | 0 | 38 | 4 | 0 |
| B19V | Galileo Analytics | 4.69 | 4.93 | 5.52 | 5.16 | 5.17 |
|  | Centrifuge | 0 | 1 | 0 | 1 | 2 |
|  | Genome Detective | 0 | 0 | 0 | 0 | 0 |
| TTV | Galileo Analytics | 5.21 |  | 5.19 | 5.25 | 5.66 |
|  | Centrifuge | 22 |  | 45 | 35 | 131 |
|  | Genome Detective | 37 |  | 168 | 97 | 547 |
| HHV6A | Galileo Analytics |  | 3.48 |  |  |  |
|  | Centrifuge |  | 1 |  |  |  |
|  | Genome Detective |  | 0 |  |  |  |
| HHV6B | Galileo Analytics |  | 4.11 |  |  |  |
|  | Centrifuge |  | 14 |  |  |  |
|  | Genome Detective |  | 14 |  |  |  |

| P4 TTV |  | S1 | S2 | S3 | S4 |
| --- | --- | --- | --- | --- | --- |
| CMV | Galileo Analytics | 2.95 | 3.46 |  |  |
|  | Centrifuge | 26 | 38 |  |  |
|  | Genome Detective | 28 | 53 |  |  |
| B19V | Galileo Analytics | 3.98 | 4.16 | 4.07 | 4.27 |
|  | Centrifuge | 2 | 1 | 1 | 3 |
|  | Genome Detective | 0 | 0 | 0 | 0 |

| P5 B19V |  | S1 | S2 | S3 |
| --- | --- | --- | --- | --- |
| CMV | Galileo Analytics | 2.71 |  | 3.46 |
|  | Centrifuge | 2 |  | 27 |
|  | Genome Detective | 4 |  | 52 |
| EBV | Galileo Analytics |  | 2.08 |  |
|  | Centrifuge |  | 0 |  |
|  | Genome Detective |  | 0 |  |
| BKV | Galileo Analytics |  |  | 3.81 |
|  | Centrifuge |  |  | 12 |
|  | Genome Detective |  |  | 20 |
| HSV1 | Galileo Analytics |  |  | 3.25 |
|  | Centrifuge |  |  | 12 |
|  | Genome Detective |  |  | 0 |
| HSV2 | Galileo Analytics |  |  | 2.76 |
|  | Centrifuge |  |  | 0 |
|  | Genome Detective |  |  | 0 |

| P6 B19V |  | S1 | S2 | S3 |
| --- | --- | --- | --- | --- |
| ADV | Galileo Analytics |  |  | 2.72 |
|  | Centrifuge |  |  | 1 |
|  | Genome Detective |  |  | 0 |
| EBV | Galileo Analytics |  | 1.19 |  |
|  | Centrifuge |  | 0 |  |
|  | Genome Detective |  | 0 |  |
| TTV | Galileo Analytics | 6.58 | 2.83 | 2.70 |
|  | Centrifuge | 1681 | 10094 | 3 |
|  | Genome Detective | 10100 | 17873 | 0 |

